# Efficacy of acupuncture and acupressure for the treatment of Raynaud’s syndrome: Protocol for a meta-analysis of randomized controlled trials

**DOI:** 10.1101/2020.06.03.20121699

**Authors:** Jiawen Deng

## Abstract

Raynaud’s syndrome is a rare vascular disorder that causes the contraction of blood vessels, usually in the fingers and toes, when there is a decrease in temperature or during emotional events. As a result, blood cannot reach the tissue in these areas, causing them to appear blue or white. It had long been speculated that acupuncture and acupressure may help mediate symptoms of Raynaud’s syndrome, however no knowledge synthesis project regarding this topic had ever been conducted (to our best knowledge). We propose a meta-analysis that investigates whether the use of acupuncture or acupressure can improve symptoms of Raynaud’s syndrome. Our proposed outcomes are incidences of positive cold water immersion test, incidences of positive temperature recovery after cold water immersion tests, incidences of remission/short-term remission, number and duration of attacks per day, and outcomes relating to nailfold microcirculation examinations (including capillary blood flow velocity, capillary deformity and capillary density).

## INTRODUCTION

Raynaud’s syndrome is a rare vascular disorder that causes the contraction of blood vessels, usually in the fingers and toes, when there is a decrease in temperature or during emotional events. As a result, blood cannot reach the tissue in these areas, causing them to appear blue or white. It had long been speculated that acupuncture and acupressure may help mediate symptoms of Raynaud’s syndrome, however no knowledge synthesis project regarding this topic had ever been conducted (to our best knowledge). We propose a meta-analysis that investigates whether the use of acupuncture or acupressure can improve symptoms of Raynaud’s syndrome.

## METHODS

We will conduct this meta-analysis in accordance to the Preferred Reporting Items for Systematic Reviews and Meta-Analyses (PRISMA) framework[1]. This study is currently being reviewed for registration on The International Prospective Register of Systematic Reviews (PROSPERO). Any significant amendments to this protocol will be reported and published with the results of the review.

### Eligibility Criteria

#### Types of Participants

We will include all patients who have been diagnosed with Raynaud’s syndrome. We will not place restrictions on age, gender or disease durations.

#### Types of Interventions

We will include all studies that include the use of any acupuncture or acupressure techniques (including, but not limited to: electroacupuncture and warm-needle acupuncture). If other treatments are used to supplement the acupuncture therapy, the treatments used must be consistent between treatment arms (i.e. acupuncture + concurrent treatment vs. concurrent treatment only).

#### Types of Studies

We will include randomized and quasi-randomized parallel-groups RCTs.

### Outcomes

#### Cold Water Immersion Test (n)

We will examine incidences of a positive cold water immersion test (onset of Raynaud’s symptoms after immersion in cold water) and incidences of temperature recovery from a cold water immersion test (temperature recovered to normal levels after a cold water test, within a predefined time period). These information should be collected at the latest follow up.

#### Incidence of Improvement or Remission (n)

We will examine incidences of symptom improvement or remission after acupuncture or acupressure, defined as per individual study criteria.

#### Number of Attacks Per Day (n±SD)

We will examine the mean number of attacks per day, collected at the latest follow up.

#### Nailfold Capillaroscopy: Number of Patients with Deformed Capillaries (n)

We will examine the number of patients with deformed capillaries, as examined via nailfold capillaroscopy, at the latest follow up.

#### Nailfold Capillaroscopy: Number of Patients with Reduced Capillary Flow (n)

We will examine the number of patients with reduced capillary flow, defined as per individual criteria. The patients should be examined using nailfold capillaroscopy at the latest follow up.

#### Nailfold Capillaroscopy: Number of Patients with Low Capillary Density (n)

We will examine the number of patients with low capillary density, as examined via nailfold capillaroscopy at the latest follow up.

### Search Methods for Identification of Studies

#### Electronic Database Search

We will conduct a database search of MEDLINE, EMBASE, Web of Science, CINAHL, and CENTRAL from inception to January 2020. We will use relevant MeSH headings to ensure appropriate inclusion of titles and abstracts (see ***Table 1*** for search strategy).

**Table 1.** MEDLINE Search Strategy

| Database: OVID Medline Epub Ahead of Print, In-Process & Other Non-Indexed Citations, Ovid MEDLINE(R) Daily and Ovid MEDLINE(R) 1946 to Present |  |
| --- | --- |
| Line |  |
| 1 | Raynaud Disease/ |
| 2 | Raynaud.ti,kf. |
| 3 | 1 or 2 |
| 4 | exp randomized controlled trial/ |
| 5 | exp Randomized Controlled Trials as Topic/ |
| 6 | randomized controlled trial.pt. |
| 7 | random*.mp. |
| 8 | double-blind method/ or single-blind method/ |
| 9 | ((singl* or doubl* or tripl* or trebl*) adj3 (blind* or mask* or procedure* or method*)).mp. |
| 10 | systematic review*.mp. |
| 11 | systematic review.pt. |
| 12 | meta analys?s.mp. |
| 13 | meta-analysis/ or "systematic review"/ |
| 14 | or/4-13 |
| 15 | 3 and 14 |

Major Chinese databases, including Wanfang Data, Wanfang Med Online, CNKI, and CQVIP will also be searched using a custom Chinese search strategy (see ***Table 2*** for search strategy).

**Table 2.** Search Strategy (CNKI)

|  |
| --- |
| ((TI='raynaud'+ 'raynauds'+ '雷诺') OR (SU='raynaud'+ 'raynauds'+ '雷诺综合征'+ '雷诺氏综合征') OR (KY='raynaud'+ 'raynauds'+ '雷诺综合征'+ '雷诺氏综合征')) AND ((TI='随机') OR (AB='随机')) |

#### Other Data Sources

We will also conduct hand search the reference list of previous meta-analyses and NMAs for included articles.

### Data Collection and Analysis

#### Study Selection

We will perform title and abstract screening independently and in duplicate using Rayyan QCRI (https://rayyan.qcri.org). Studies will only be selected for full-text screening if both reviewers deem the study relevant. Full-text screening will also be conducted in duplicate. We will resolve any conflicts via discussion and consensus or by recruiting a third author for arbitration.

#### Data Collection

We will carry out data collection independently and in duplicate using data extraction sheets developed a priori. We will resolve discrepancies by recruiting a third author to review the data.

#### Risk of Bias

We will assess risk of bias (RoB) independently and in duplicate using The Cochrane Collaboration’s tool for assessing risk of bias in randomized trials[2]. Two reviewers will assess biases within each article in seven domains: random sequence generation, allocation concealment, blinding of participants and personnel, blinding of outcome assessment, incomplete outcome data, selective reporting, and other biases.

If a majority of domains are considered to be low risk, the study will be assigned a low RoB. Similarly, if a majority of domains are considered to be high risk, the study will be assigned a high RoB. If more than half of the domains have unclear risk or if there are an equivalent number of low and high, low and unclear or high and unclear domains, the study will be assigned an unclear RoB.

### Data Items

#### Bibliometric Data

Authors, year of publication, trial registration, digital object identifier (DOI), publication journal, funding sources and conflict of interest.

#### Methodology

\# of participating centers, study setting, blinding methods, phase of study, enrollment duration, randomization and allocation methods, criteria for remission.

#### Baseline Data

\# randomized, # analyzed, mean age, sex, follow up duration.

#### Outcome Related Data

Please see the **Outcomes** section.

### Statistical Analysis

#### Meta-Analysis

We will conduct all statistical analyses using R 4.0.0[3]. We will perform meta-analyses using the *meta* library. Because we expect significant heterogeneity among studies due to differences in methodology, we will use a random effects model[4].

For dichotomous outcomes, we will report the results of the analyses as risk ratio (RR), pooled using the Mantel-Haenszel method[5], with 95% confidence intervals (CIs) with Hartung-Knapp adjustment for random effects model[6]. For continuous outcomes, we will report the results as mean differences (MDs), pooled using the inverse variance method[7], with corresponding 95% CIs with Hartung-Knapp adjustment for random effects model[6]. We used the Sidik-Jonkman estimator[6,7] for τ^2^ calculations, and we used the Q-profile method[8] for estimating the confidence interval of τ^2^ and τ.

#### Missing Data

We will attempt to contact the authors of the original studies to obtain missing or unpublished data. We will attempt to impute standard deviation values if not provided by the study.

#### Heterogeneity Assessment

We will assess statistical heterogeneity using I^2^ statistics, τ^2^ and Cochran’s Q[9]. We will identify the source of heterogeneity by

a. identify outlier studies with treatment effect that is not included in the 95% confidence interval of the pooled effect size using the *dmetar* library;
b. perform influence analyses, or the “leave-one-out” analysis, where the meta-analysis is repeated with one study omitted, using the *dmetar* library;
c. Perform GOSH analyses[10] using the *metafor* library. The GOSH plot will be examined for evaluable clusters, and *gosh.diagnostics* function will be used to identify the outlying studies if there are evaluable clusters.

We will perform sensitivity analyses excluding outlier studies to observe the outliers’ effects on the original pooled effect size and heterogeneity measures.

#### Publication Bias

We will use funnel plots[11] to detect the presence of small study effects. We will use Egger’s test[12] to check for asymmetry within the funnel plot to identify possible publication bias. If Egger’s test reveals significant publication bias, we will use the trim-and-fill method[13] to estimate the actual effect size with imputations of the missing small studies. This will be done using the *trimfill* method in the *meta* library.

We will also perform p-curve [14] analyses to detect the presence of “p-hacking” [15] using the *dmetar* library. We will report whether we observed evidence of “p-hacking”, such as a lack of right skew in the p-curve plot or low estimated statistical power.

#### Meta-Regression

We will perform meta-regression on:

1. % of patients with primary/secondary Raynaud’s syndrome
2. gender (% female patients)
3. follow up periods

## Data Availability

No data available.

## ACKNOWLEDGEMENTS

None

## AUTHOR STATEMENT

JD made significant contributions to conception and design of the work, drafted the work, and substantially reviewed it.

## REFERENCES

1 Moher D, Liberati A, Tetzlaff J, et al. Preferred reporting items for systematic reviews and meta-analyses: the PRISMA statement. Ann Intern Med 2009; 151:264–9, W64. doi:10.7326/0003-4819-151-4-200908180-00135

2 Higgins JPT, Altman DG, Gotzsche PC, et al. The Cochrane Collaboration’s tool for assessing risk of bias in randomised trials. BMJ. 2011;343:d5928–d5928. doi:10.1136/bmj.d5928

3 Team RC. R: A language and environment for statistical computing. 2015.

4 Serghiou S, Goodman SN. Random-Effects Meta-analysis: Summarizing Evidence With Caveats. JAMA 2019;321:301–2. doi:10.1001/jama.2018.19684

5 Mathes T, Kuss O. A comparison of methods for meta-analysis of a small number of studies with binary outcomes. Res Synth Methods 2018;9:366–81. doi:10.1002/jrsm.1296

6 IntHout J, Ioannidis JPA, Borm GF. The Hartung-Knapp-Sidik-Jonkman method for random effects meta-analysis is straightforward and considerably outperforms the standard DerSimonian-Laird method. BMC Med Res Methodol 2014;14:25. doi:10.1186/1471-2288-14-25

7 Marín-Martínez F, Sánchez-Meca J. Weighting by Inverse Variance or by Sample Size in Random-Effects Meta-Analysis. Educ Psychol Meas 2010;70:56–73. doi:10.1177/0013164409344534

8 Jackson D, Turner R, Rhodes K, et al. Methods for calculating confidence and credible intervals for the residual between-study variance in random effects meta-regression models. BMC Med Res Methodol 2014;14:103. doi:10.1186/1471-2288-14-103

9 Higgins JPT, Thompson SG, Deeks JJ, et al. Measuring inconsistency in meta-analyses. BMJ 2003;327:557–60. doi:10.1136/bmj.327.7414.557

10 Olkin I, Dahabreh IJ, Trikalinos TA. GOSH – a graphical display of study heterogeneity. Res Synth Methods 2012;3:214–23. doi:10.1002/jrsm.1053

11 Sterne JAC, Sutton AJ, Ioannidis JPA, et al. Recommendations for examining and interpreting funnel plot asymmetry in meta-analyses of randomised controlled trials. BMJ 2011;343:d4002. doi:10.1136/bmj.d4002

12 Lin L, Chu H. Quantifying publication bias in meta-analysis. Biometrics 2018;74:785–94. doi:10.1111/biom.12817

13 Duval S, Tweedie R. Trim and fill: A simple funnel-plot-based method of testing and adjusting for publication bias in meta-analysis. Biometrics 2000;56:455–63. doi:10.1111/j.0006-341x.2000.00455.x

14 Simonsohn U, Nelson LD, Simmons JP. P-curve won’t do your laundry, but it will distinguish replicable from non-replicable findings in observational research: Comment on Bruns & Ioannidis (2016). PLoS One. 2019;14:e0213454. doi:10.1371/journal.pone.0213454

15 Head ML, Holman L, Lanfear R, et al. The extent and consequences of p-hacking in science. PLoS Biol 2015;13:e1002106. doi:10.1371/journal.pbio.1002106

